# Potential biochemical markers to identify severe cases among COVID-19 patients

**DOI:** 10.1101/2020.03.19.20034447

**Authors:** Jialin Xiang, Jing Wen, Xiaoqing Yuan, Shun Xiong, Xue Zhou, Changjin Liu, Xun Min

**Affiliations:** Department of Laboratory Medicine, Affiliated Hospital of Zunyi Medical University, Guizhou, China; Department of Medical Imaging, Affiliated Hospital of Zunyi Medical University, Guizhou, China; Department of Laboratory Medicine, The Fourth People’s Hospital of Zunyi city, Guizhou, China; School of Laboratory Medicine, Zunyi Medical University, Guizhou, China

**Keywords:** Severe COVID-19, diagnosis, serum biomarkers, glomerular filtration function injury

## Abstract

There is a high mortality and long hospitalization period for severe cases with 2019 novel coronavirus disease (COVID-19) pneumonia. Therefore, it makes sense to search for a potential biomarker that could rapidly and effectively identify severe cases early. Clinical samples from 28 cases of COVID-19 (8 severe cases, 20 mild cases) in Zunyi District from January 29, 2020 to February 21, 2020 were collected and otherwise statistically analysed for biochemical markers. Serum urea, creatinine (CREA) and cystatin C (CysC) concentrations in severe COVID-19 patients were significantly higher than those in mild COVID-19 patients (P<0.001), and there were also significant differences in serum direct bilirubin (DBIL), cholinesterase (CHE) and lactate dehydrogenase (LDH) concentrations between severe and mild COVID-19 patients (P<0.05). Serum urea, CREA, CysC, DBIL, CHE and LDH could be used to distinguish severe COVID-19 cases from mild COVID-19 cases. In particular, serum biomarkers, including urea, CREA, CysC, which reflect glomerular filtration function, may have some significance as potential indicators for the early diagnosis of severe COVID-19 and to distinguish it from mild COVID-19. Glomerular filtration function injury in severe COVID-19 patients should also be considered by clinicians.

## Introduction

To date, the 2019 novel coronavirus pneumonia (COVID-19) is the greatest public health problem in the world. By 10:30 on Mar 19, 2020, the cumulative number of confirmed cases had reached 218,785 globally, including 8,949 deaths. Of the total confirmed COVID-19 patients, about 19.9% were severe cases[1], which have a mortality rate of about 20%. Severe cases often suffer from underlying diseases such as cardiovascular disease and diabetes that can accelerate the progression of 2019 novel coronavirus (SARS-CoV-2) infection[2, 3]. Furthermore, acute respiratory distress syndrome could lead to death in some severe COVID-19 patients, and this is often accompanied by heart failure, liver failure, and kidney failure[2, 4]. Therefore, early, simple and effective diagnosis of severe COVID-19 pneumonia is of great significance in reducing mortality and shortening the hospitalization period.

The guidelines on the diagnosis and treatment of novel coronavirus pneumonia[5-7] have clear criteria for severe COVID-19, including respiratory rate, haemoglobin oxygen saturation (SaO2), oxygenation index (PaO2/FiO2), and so on. However, these criteria are susceptible to subjective and objective factors, which may lead to an extended time for diagnosing and the possibility of misdiagnosing severe COVID-19. Therefore, it makes sense to find a potential biomarker that could effectively diagnose severe COVID-19.

In this study, we investigated and analysed more than 4,000 test results from 28 COVID-19 patients. The concentrations of 6 serum biochemical markers showed significant differences between severe and mild COVID-19 patients. Interestingly, the levels of serum biomarkers reflecting glomerular filtration function (including cystatin C (CysC), creatinine (CREA), and urea) in severe COVID-19 patients were significantly higher than those in mild patients. It has been demonstrated that serum urea, CREA and CysC could be potential biomarkers that could rapidly and specifically reflect severe COVID-19, and it is of great significance in the early diagnosis of COVID-19 to reduce the mortality and shorten the hospitalization period for patients with severe COVID-19. Moreover, glomerular filtration injury in patients with severe COVID-19 should not be ignored by clinicians.

## Materials and Methods

The study enrolled patients who were diagnosed with COVID-19 at The First Affiliated Hospital of Zunyi Medical University and The Fourth People’s Hospital of Zunyi city from January 29, 2020 to February 21, 2020. The exclusion criteria were based on the guidelines for the diagnosis and treatment of novel coronavirus pneumonia[6]. The inclusion criteria for participants are shown in Figure 1.

**Figure 1.**
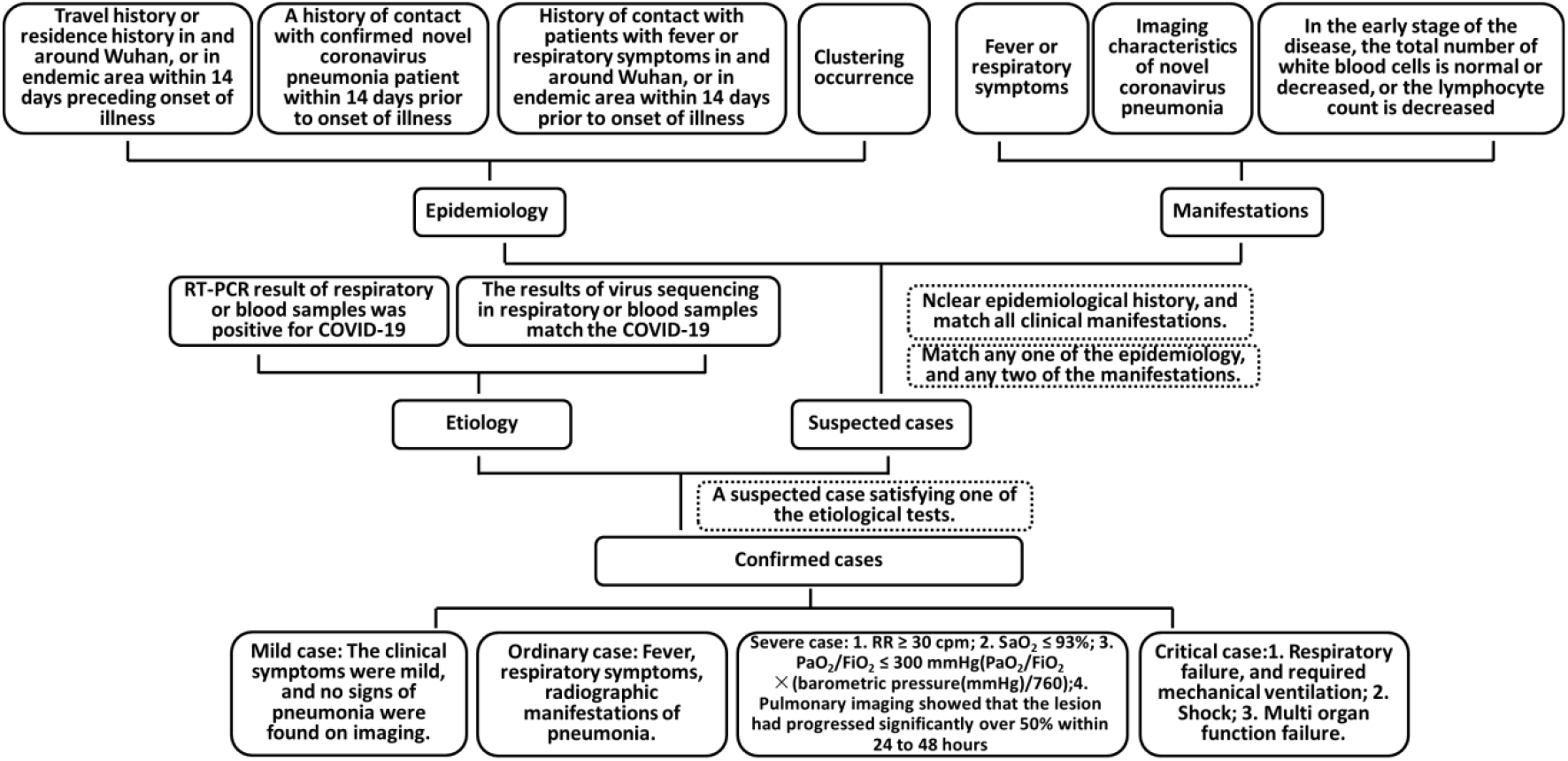
Chart representing the guidelines for the diagnosis of novel coronavirus pneumonia.

The analysis included 4,166 test results from 28 COVID-19 patients, including 8 confirmed cases of severe COVID-19 (with a mean age of 66±22 years) and 20 confirmed cases of mild COVID-19 (with a mean age of 41±19 years), and the remaining baseline characteristics are shown in the Table 1. The serum biochemical analysis data, including aspartate aminotransferase (AST), glutamyl transferase (GGT), alkaline phosphatase (ALP), cholinesterase (CHE), total bilirubin (TBIL), direct bilirubin (DBIL), creatine kinase (CK), creatine phosphokinase-isoenzyme-MB (CKMB), lactate dehydrogenase (LDH), hydroxybutyrate dehydrogenase (HBDH), CysC, CREA, urea, uric acid (UA), bicarbonate (HCO^3-^) and C-reaction protein (CRP), were collected from COVID-19 patients when routine blood screening tests were performed. The above biomarkers were all determined by detection kits (Beckman Coulter, Suzhou, China) on a Beckman Coulter AU5821 biochemical analyser (Tokyo, Japan). Low- and high-value human serum matrix standards from Bio-Rad (California, USA) were used as quality controls in the biomarker tests, and all biochemical indexes were approved by the National Center for Clinical Laboratories (NCCL) external quality assessment programmes in laboratory medicine.

**Table 1.** Baseline characteristics of the study subjects.

|  | Severe cases | Mild cases |
| --- | --- | --- |
| N | 8 | 20 |
| Gender (male/female) | 4/4 | 11/9 |
| Age (years) | 66 ± 22 | 41 ± 19 |
| Diabetes mellitus (N, %) | 2, 25.0% | 2, 10.0% |
| Hypertension (N, %) | 4, 50.0% | 1, 5.0% |
| Cardiovascular disease (N, %) | 1, 12.5% | 3, 15.0% |
| Nephropathy (N, %) | 0, 0% | 1, 5.0% |

The statistical analysis was performed using IBM SPSS Statistics 22.0 (New York, USA) software. The Wilcoxon signed-rank test was used to compare the data between two groups. Differences with a value of P<0.05 were considered statistically significant.

## Result

In this study, the majority of serum biochemical markers, including UA, HCO^3-^, AST, GGT, ALP, TBIL, TBA, TP, ALB, PA, HBDH, CK, CKMB and CRP, did not show significant differences between severe and mild COVID-19 patients. The differences in and the dispersion of the biomarker results between severe and normal COVID-19 patients are shown in Figure 2.

**Figure 2.**
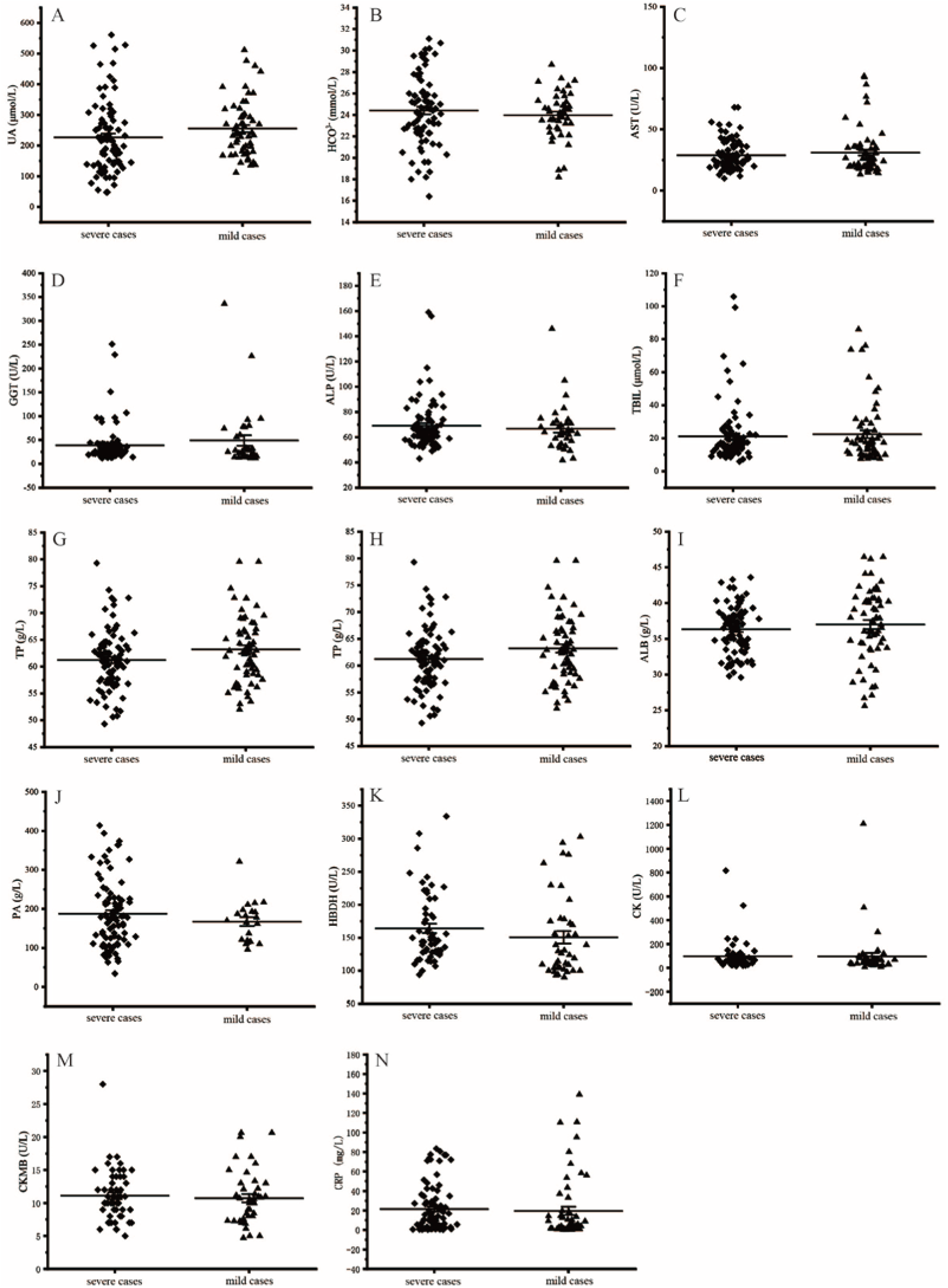
Distributions showing serum biochemical marker levels in severe and mild COVID-19 cases.

Glomerular filtration function markers, such as serum urea, CREA and CysC, were significantly higher in severe COVID-19 patients than in mild COVID-19 patients (*P*<0.001) (Figure 3A, B, C). There were also significant differences in serum DBIL, CHE and LDH concentrations between severe and mild COVID-19 patients (*P*<0.05) (Figure 3D, E, F).

**Figure 3.**
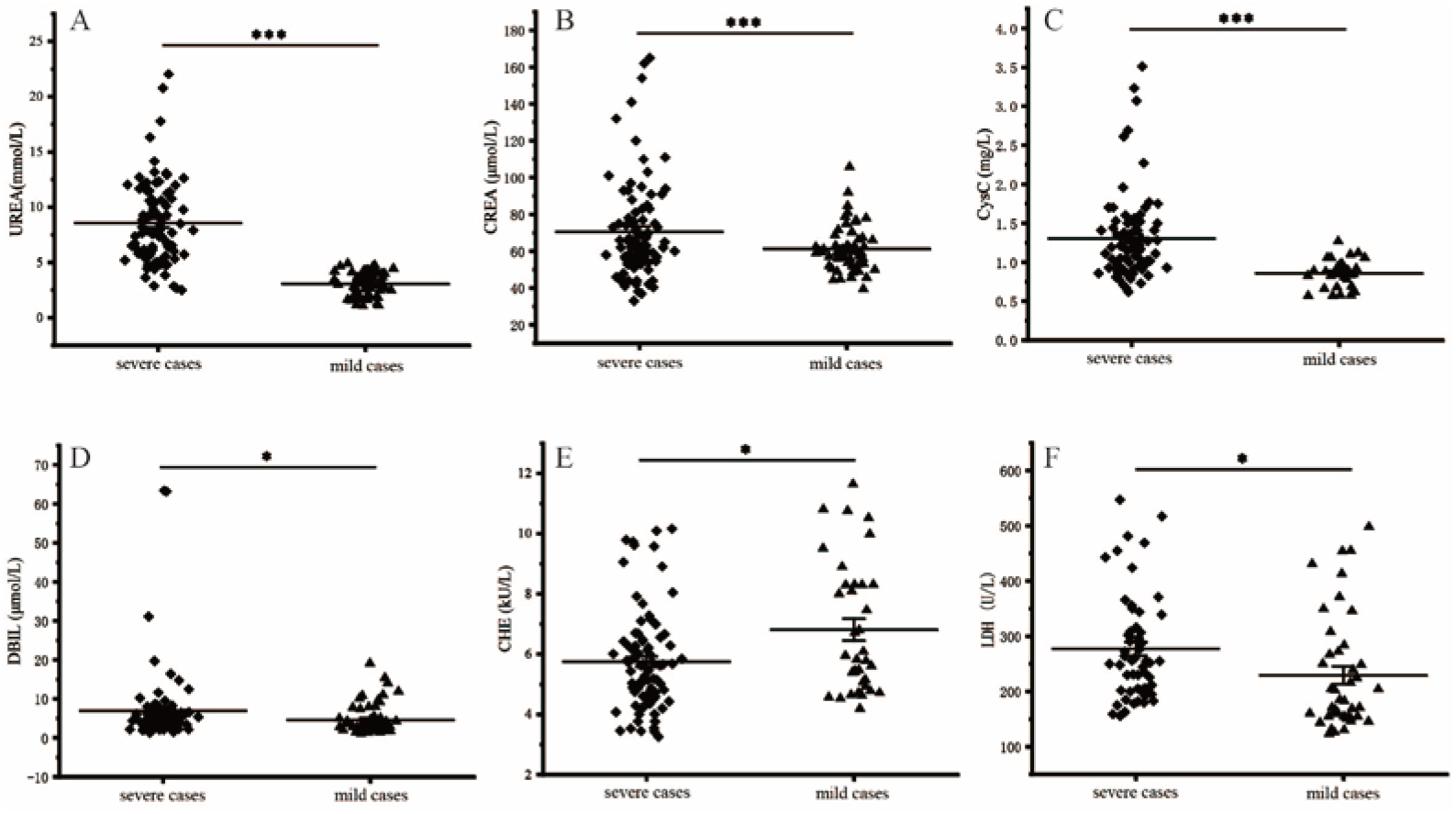
Distributions and differences showing the concentration of serum urea, CREA, CysC, DBIL, CHE, LDH between severe and mild COVID-19 cases. ***P<0.001, *P<0.05.

## Discussion

In this paper, the results of most serum biochemical tests were investigated and analysed. The incidence of liver or heart failure was higher in severe COVID-19 patients[2, 4]; however, we found that most of the indicators, including liver and cardiac function biomarkers, except for serum DBIL, CHE and LDH, did not change significantly or directly in severe and mild patients. There were not much consistent with some reports in the literature[2], which may be the result of prophylactic use of drug by doctors. There were also no differences in serum CRP concentrations between severe and mild COVID-19 patients; however, the overall level of serum CRP increased, suggesting that it could be sensitive to SARS-CoV-2 infection.

The results of renal injury were similar with 2003 SARS-CoV[8], a close relative of SARS-CoV-2. We also found that serum urea, CREA, and CysC, which are biochemical markers of renal function, were significantly elevated in severe COVID-19 patients, suggesting that SARS-CoV-2 infection could damage the kidney, and this is consistent with part of the latest findings[9]. Furthermore, there were no differences in the serum HCO^3-^ results between severe and mild COVID-19 patients, and this further suggests that the glomerulus could be one of the target organs for the coronavirus in severe patients and may be related to the high expression of ACE2 in glomerular cells[10, 11].

Two limitations are inherent in our study. First, the survey area is relatively limited. Additionally, the relatively small number of participants in the survey may not fully reflect the overall situation. Thus, further studies are necessary to execute a large number of data surveys in multiple regions by multi-centre cooperation.

Herein, we found that some serum indicators of glomerular function, including urea, CREA, and CysC, were observably higher in severe COVID-19 patients than in mild COVID-19 patients. This result indicates that serum urea, CREA, and CysC could be used as potential biochemical biomarkers to identify a severe COVID-19 patient. Moreover, clinicians should constantly monitor the glomerular filtration function while treating patients with a severe SARS-CoV-2 infection.

## Data Availability

All data, models, and code generated or used during the study appear in the submitted article.

## Acknowledgements

The authors wish to thank all the patients who enrolled in this study.

## Ethical Statement

This study was approved by the Ethics Commissions of the Affiliated Hospital of Zunyi Medical University (No. KLL-2020-008).

## References

[1] Report of the CHINA-WHO joint investigation on new coronavirus pneumonia (COVID - 19), http://www.nhc.gov.cn/jkj/s3578/202002/87fd92510d094e4b9bad597608f5cc2c/files/e73a238d8eff45d5ab855fa078f4c0dd.pdf, (2020) (accessed 29 February 2020)..

[2] C. Huang, Y. Wang, X. Li, L. Ren, J. Zhao, Y. Hu, L. Zhang, G. Fan, J. Xu, X. Gu, Z. Cheng, T. Yu, J. Xia, Y. Wei, W. Wu, X. Xie, W. Yin, H. Li, M. Liu, Y. Xiao, H. Gao, L. Guo, J. Xie, G. Wang, R. Jiang, Z. Gao, Q. Jin, J. Wang, B. Cao, Clinical features of patients infected with 2019 novel coronavirus in Wuhan, China, Lancet (London, England), 395 (2020) 497–506.

[3] C. N, Z. M, D. X, Q. J, G. F, H. Y, Q. Y, W. J, L. Y, W. Y, X. J, Y. T, Z. X, Z. L, Epidemiological and clinical characteristics of 99 cases of 2019 novel coronavirus pneumonia in Wuhan, China: a descriptive study, Lancet (London, England), 395 (2020) 507–513.

[4] D. Wang, B. Hu, C. Hu, F. Zhu, X. Liu, J. Zhang, B. Wang, H. Xiang, Z. Cheng, Y. Xiong, Y. Zhao, Y. Li, X. Wang, Z. Peng, Clinical Characteristics of 138 Hospitalized Patients With 2019 Novel Coronavirus–Infected Pneumonia in Wuhan, China, JAMA, (2020).

[5] N.H.C.o.t.P.s.R.o. China., The notice of launching guideline on diagnosis and treatment of the novel coronavirus pneumonia (NCP). Revised version of the 5th edition., (2020).

[6] N.H.C.o.t.P.s.R.o. China, The notice of launching guideline on diagnosis and treatment of the novel coronavirus pneumonia (NCP). 6th edition., (2020).

[7] N.H.C.o.t.P.s.R.o. China, The notice of launching guideline on diagnosis and treatment of the novel coronavirus pneumonia (NCP). 5th edition., (2020).

[8] C. Kh, T. Wk, T. Cs, L. Mf, L. Fm, T. Kf, F. Ks, T. Hl, Y. Ww, C. Hw, L. Ts, T. Kl, L. Kn, Acute renal impairment in coronavirus-associated severe acute respiratory syndrome, Kidney international, 67 (2005) 698–705.

[9] Z. Li, M. Wu, J. Guo, J. Yao, X. Liao, S. Song, M. Han, J. Li, G. Duan, Y. Zhou, X. Wu, Z. Zhou, T. Wang, M. Hu, X. Chen, Y. Fu, C. Lei, H. Dong, Y. Zhou, H. Jia, X. Chen, J. Yan, Caution on Kidney Dysfunctions of 2019-nCoV Patients, medRxiv, (2020) 2020.2002.2008.20021212.

[10] Z. P, Y. XL, W. XG, H. B, Z. L, Z. W, S. HR, Z. Y, L. B, H. CL, C. HD, C. J, L. Y, G. H, J. RD, L. MQ, C. Y, S. XR, W. X, Z. XS, Z. K, C. QJ, D. F, L. LL, Y. B, Z. FX, W. YY, X. GF, S. ZL, A pneumonia outbreak associated with a new coronavirus of probable bat origin, Nature, (2020).

[11] L. W, M. MJ, V. N, S. J, W. SK, B. MA, S. M, S. JL, L. K, G. TC, C. H, F. M, Angiotensin-converting enzyme 2 is a functional receptor for the SARS coronavirus, Nature, 426 (2003) 450–454.

